# An Early Return-to-Work Program for COVID-19 Close Contacts in Healthcare During the Omicron Wave in Japan

**DOI:** 10.1101/2022.05.02.22274478

**Authors:** Hiroyuki Naruse, Yohei Doi, Mitsunaga Iwata, Kiyohito Ishikawa

**Affiliations:** Department of Clinical Pathophysiology, Fujita Health University School of Medical Sciences, 1-98 Kutsukake-cho, Dengakugakubo, Toyoake 470-1192, Japan; Department of Infectious Diseases, Fujita Health University School of Medicine, 1-98 Kutsukake-cho, Dengakugakubo, Toyoake 470-1192, Japan; Department of Emergency Medicine and General Internal Medicine, Fujita Health University School of Medicine, 1-98 Kutsukake-cho, Dengakugakubo, Toyoake 470 -1192, Japan; Department of Quality and Safety in Healthcare, Division of Infection Control and Prevention, Fujita Health University Hospital, 1-98 Kutsukake-cho, Dengakugakubo, Toyoake 470-1192, Japan

**Keywords:** COVID-19, Return-to-Work Program, Healthcare workers

## Abstract

During the coronavirus disease 2019 (COVID-19) pandemic, maintaining adequate staffing in healthcare facilities is important to provide a safe work environment for healthcare workers (HCWs). Japan’s early return-to-work (RTW) program may be a rational strategy at a time when there is an increased demand for the services of HCWs. We assessed whether the early RTW program for HCWs who have been in close contact with a COVID-19 case in our hospital was justified. Close contacts were identified according to the guidance of the World Health Organization. Between January and March 2022, 256 HCWs were identified as close contacts (median age, 35 years; 192 female). Thirty-seven (14%) secondary attack cases of COVID-19 were detected. Among 141 HCWs who applied to the early RTW program, nurses and doctors comprised about three-quarters of participants, with a higher participation rate by doctors (78%) than nurses (59%). Eighteen HCWs tested positive for COVID-19 by the sixth day after starting the early RTW program. No COVID-19 infection clusters were reported during the observation period. These findings suggest that the early RTW program for COVID-19 close contacts was a reasonable strategy for HCWs during the Omicron wave.

## Introduction

Healthcare workers (HCWs) are vulnerable to infection due to frequent and close contact with coronavirus disease 2019 (COVID-19) patients (1,2). Quarantining close contacts of confirmed cases is a universal strategy to prevent onward transmission of the severe acute respiratory syndrome coronavirus 2 (SARS-CoV-2) (3). During the COVID-19 pandemic, maintaining adequate staffing in healthcare facilities can help ensure a safe work environment for HCWs. Following the spread of infection with the SARS-CoV-2 Omicron variant, Japan’s Ministry of Health, Labour and Welfare notified to the public that HCWs who had been in contact with COVID-19 cases were able to continue working under certain conditions. However, this “early return-to-work (RTW) strategy” may increase the risk of onward transmission in the healthcare environment, including to patients who may be at an elevated risk of severe COVID-19. In this study, we assessed whether the early RTW program for COVID-19 close contacts in our hospital was justified.

## Methods

### Study design and participants

This study was a process evaluation of our routine contact tracing program at Fujita Health University Hospital (Toyoake, Japan), a 1435-bed tertiary-care academic medical center. Data were collected from January to March 2022. HCWs who were close contacts of confirmed COVID-19 cases and then quarantined were enrolled in the study. The ethics committee of Fujita Health University approved this study (study protocol number 21-400).

### Definitions

Close contacts were identified according to the guidance of the World Health Organization (4). The criteria used to differentiate “close” from “non-close” contacts comprised distance, time, and personal protective equipment (PPE) used. A cumulative total of 15 min or longer over a 24-hour period were deemed “close” if they occurred within 1 m of the index case in the absence of appropriate PPE within 2 days before their symptom onset or sample collection. “Household contacts” were defined as individuals who shared the same residential address as the index case, regardless of duration or proximity of contact.

The health management office directed the concerned departments to provide a list of all HCWs who may have come into contact with confirmed COVID-19 cases when a new index COVID-19 case was identified. Upon obtaining the list of COVID-19-exposed HCWs, Health Management office staff contacted each HCW by phone to elicit histories related to exposure durations and types, the procedures performed on the patient, and the use of PPE.

### RTW programs

The RTW program for COVID-19 close contacts in our hospital is presented in Fig.

1. All close contact HCWs were tested once using reverse transcription polymerase chain reaction (RT-PCR) tests for COVID-19 regardless of their symptoms. If the initial COVID-19 test result was negative, eligible asymptomatic HCWs were enrolled in each RTW program. If HCWs could quarantine at home or in institution-designated facilities, then early or standard RTW programs were selected. Among them, HCWs who met all of the following conditions were eligible to apply to an early RTW program: (1) difficult to replace with another HCW, (2) received the third dose of a COVID-19 mRNA vaccine at least a week earlier, (3) a negative COVID-19 antigen test before each work shift, and (4) consent from relevant HCWs and managers to participate in the program. A close contact was allowed to end their quarantine and return to work six days after their last exposure if the second COVID-19 RT-PCR test was negative. If HCWs were not able to quarantine, then they were asked to complete late or very late RTW programs depending on their home and work conditions. During the quarantine period, close contact HCWs were required to undergo another COVID-19 RT-PCR test if they showed any relevant symptoms (fever, cough, or other respiratory symptoms).

### Measurements

The COVID-19 RT-PCR and antigen tests were performed with nasopharyngeal swabs using the GENECUBE HQ SARS-CoV-2 system (TOYOBO CO., LTD, Japan) and Quick Navi COVID19Ag (Denka Company Limited, Japan), respectively.

### Results

The baseline characteristics of the study participants are summarized in Table 1. Our analysis included data related to 211 COVID-19 cases reported during the study period. In connection with these cases, 256 HCWs were identified as close contacts. The median (interquartile range) age was 35 (25–43) years and 192 of these HCWs were female. All HCWs participated in the RTW program; 141 participated in the early RTW program, 59 in the standard RTW program, 23 in the late RTW program, and 33 in the very late RTW program (Fig. 1). Among the 256 close contacts, 37 (14%) secondary attack cases of COVID-19 were detected.

**Table 1.** Baseline characteristics of the study participants according to return-to-work program.

|  | ***All***<br>***(n=256)*** | ***Early RTW***<br>***(n=141)*** | ***Standard RTW***<br>***(n=59)*** | ***Late RTW***<br>***(n=23)*** | ***Very late RTW***<br>***(n=33)*** |
| --- | --- | --- | --- | --- | --- |
| ***Age, years*** | 35 (25-43) | 32 (25-40) | 28 (24-41) | 40 (36-45) | 44 (36-50) |
| ***Female, n (%)*** | 192 (75) | 103 (73) | 47 (80) | 16 (70) | 26 (79) |
| ***Job category, n (%)*** |  |  |  |  |  |
| ***Nurse*** | 119 (47) | 70 (50) | 28 (47) | 10 (44) | 11(33) |
| ***Doctor*** | 46 (18) | 36 (26) | 3 (5) | 5 (22) | 2 (6) |
| ***Medical clerk*** | 44 (17) | 12 (9) | 17 (29) | 4 (17) | 11 (33) |
| ***Other health care professional*** | 47 (18) | 23 (16) | 11 (19) | 4 (17) | 9 (27) |
| ***Exposure setting, n (%)*** |  |  |  |  |  |
| ***Household*** | 173 (68) | 85 (60) | 32 (54) | 23 (100) | 33 (100) |
| ***Private*** | 12 (5) | 6 (4) | 6 (10) | 0 (0) | 0 (0) |
| ***In-hospital (HCWs)*** | 40 (16) | 24 (17) | 16 (27) | 0 (0) | 0 (0) |
| ***In-hospital (patients)*** | 31 (12) | 26 (18) | 5 (8) | 0 (0) | 0 (0) |
| ***Secondary attack rate, n (%)*** | 37 (14) | 18 (13) | 4 (7) | 9 (39) | 6 (18) |
| ***Time from starting program to positive results, day (develop symptoms)*** |  |  |  |  |  |
| -3 | 13 (6) | 8 (2) | 1 (1) | 3 (2) | 1 (1) |
| 4-6 | 19 (5) | 10 (1) | 3 (1) | 4 (1) | 2 (2) |
| 7- | 5 (0) | 0 (0) | 0 (0) | 2 (0) | 3 (0) |

**Figure 1.**
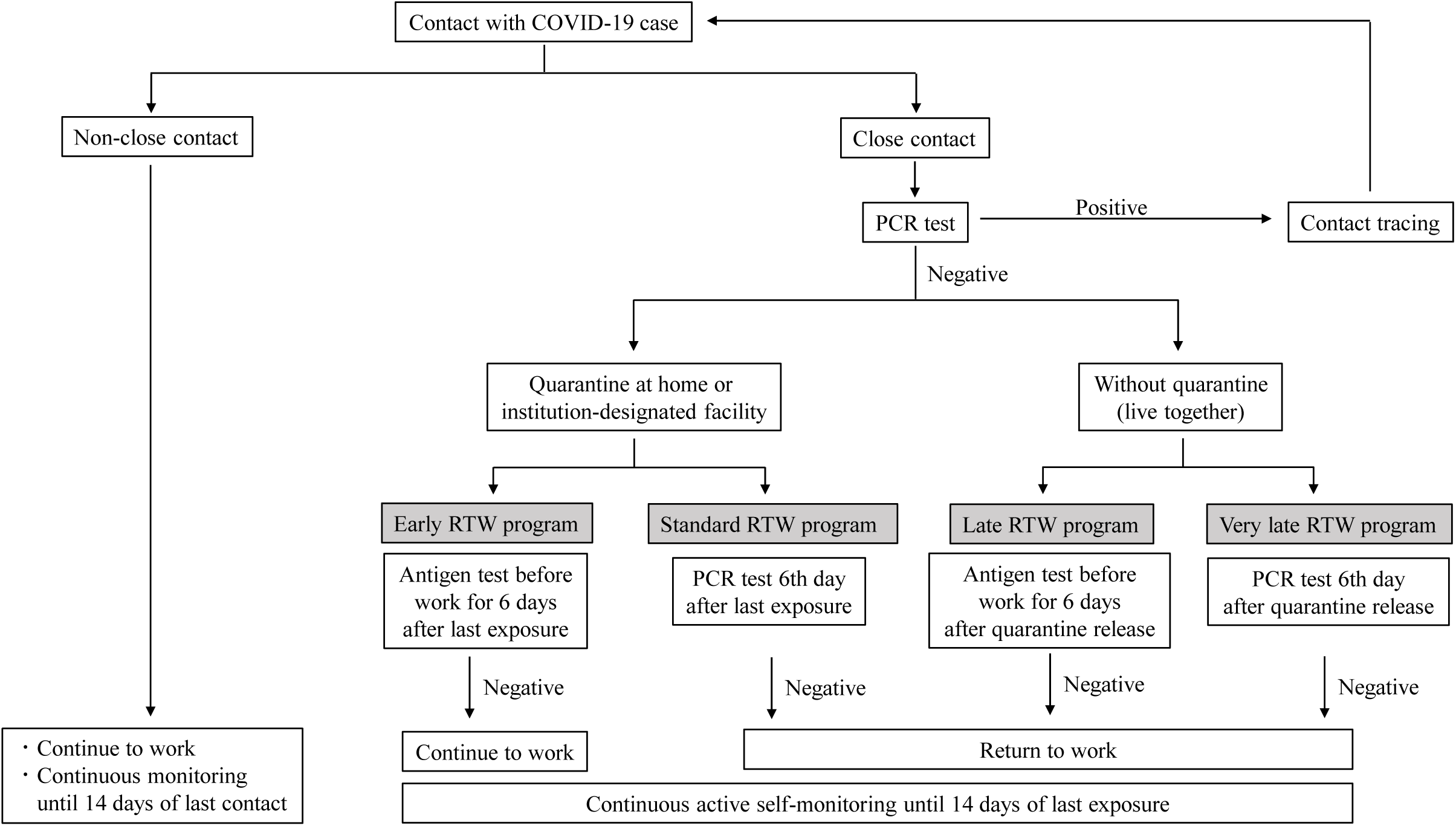
Return-to-work program for close contact HCWs in Fujita Health University Hospital

Among the 141 HCWs who participated in the early RTW program, 70 (50%) were nurses, 36 (26%) were doctors, 23 (16%) were other healthcare professionals, and 12 (9%) were medical clerks (Table 1). Doctors most frequently participated in the early RTW program (78%), followed by nurses (59%), other health care professionals (49%), and medical clerks (27%) (Fig. 2). Eighteen HCWs tested positive for a SARS-CoV-2 infection within six days of starting the early RTW program; eight participants tested positive three days (two of these HCWs developed symptoms) and ten participants tested positive between the fourth day and the sixth day (one developed symptoms). No participants tested positive after the seventh day of the program. No COVID-19 infection clusters were reported during the observation period.

**Figure 2.**
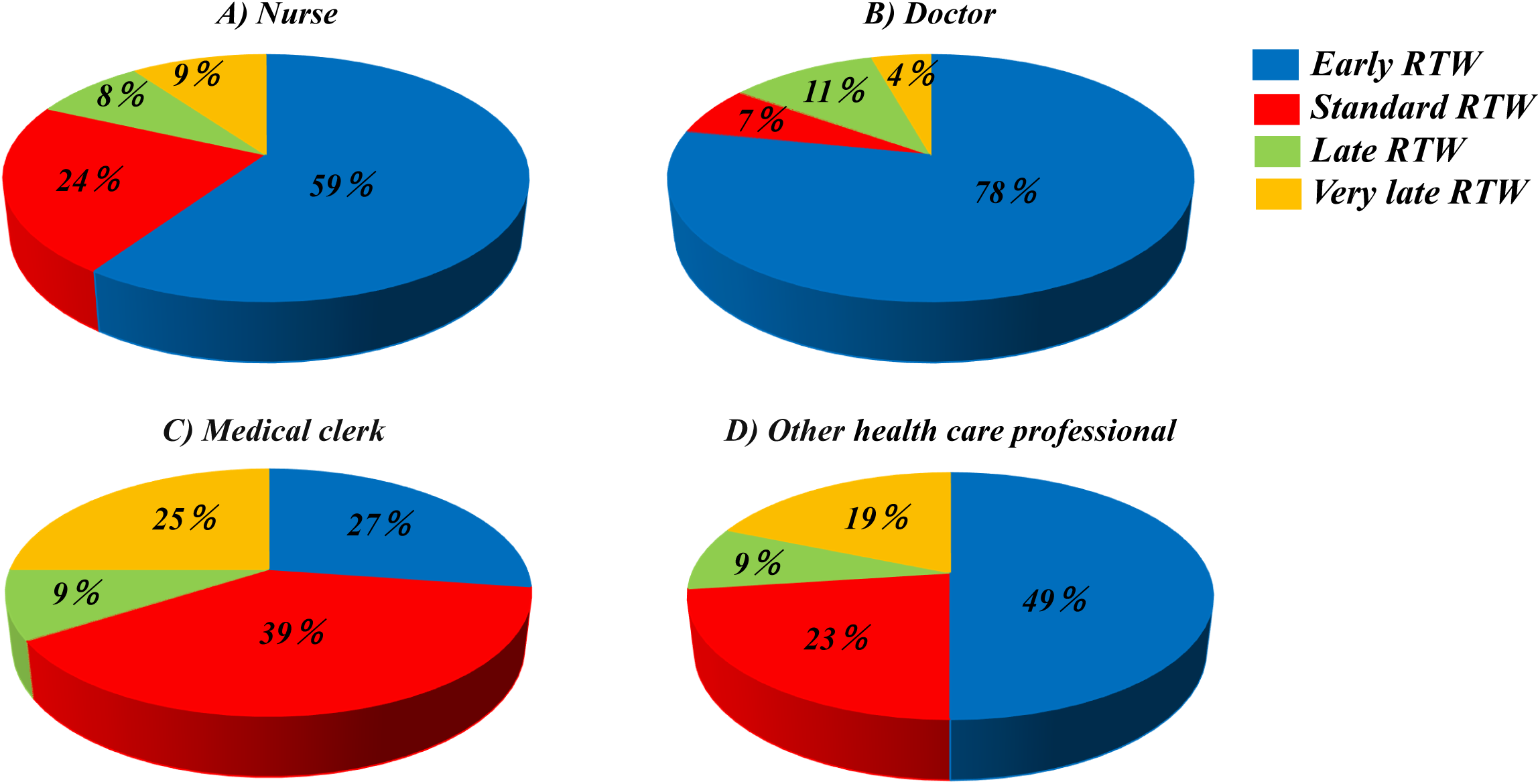
Return-to-work program according to job category

### Discussion

In total, 55% of HCWs who were close contacts of a COVID-19 case participated in the early RTW program. All participants were able to safely complete the program without infection clusters. These results suggest that an early RTW program may ameliorate workforce shortages; accordingly, it may be an especially useful strategy given the rising demand for HCWs during the COVID-19 pandemic.

Our data showed 14% of HCWs had a secondary attack, which was consistent with previous studies (5,6). Sahoo et al. reported that 3.8% of HCWs with high-risk contacts tested positive for COVID-19. Other studies have emphasized that SARS-CoV-2 is more transmissible within households compared to other contact settings (7,8)— more than half of the cases (68%) in this study had been in contact with someone in their household who had tested positive for COVID-19. Differences in exposure settings may affect the results.

A prospective study including 2761 close contacts observed no secondary attack cases of COVID-19 among participants initially exposed to the index case after six days of symptom onset (9). Benea et al. reported that all secondary attack cases of COVID-19 infection among HCWs following high-risk exposures to SARS-CoV-2 were diagnosed within six days of the last exposure (5). The National Institute of Infectious Diseases demonstrated that the number of from which live virus was recovered cases decreased six days after diagnosis among asymptomatic COVID-19 patients (10). Therefore, our early RTW program, with its six-day examination period, may be considered reasonable.

Our study has limitations. The early RTW program was launched during the Omicron wave; it is uncertain whether the program may be appropriate when other or new variants are predominant. Nurses and doctors working at a tertiary care setting comprised about three-quarters of the early RTW program participants; it is therefore necessary to consider whether this program can be extrapolated to other professions and work environments. Additionally, early RTW programs require sufficient medical resources, such as rapid diagnostic testing.

## Conclusion

In conclusion, this study suggests that early RTW programs for COVID-19 close contacts may be appropriate for HCWs during Omicron waves.

## Data Availability

All data produced in the present study are available upon reasonable request to the authors

## Acknowledgments

The authors thank the staff at the Health Management Office, COVID-19 Countermeasures Headquarters, and Division of Infection Control and Prevention in Fujita Health University Hospital.

